# Human Papilloma Virus Circulating Cell-Free DNA Kinetics in Cervical Cancer Patients Undergoing Definitive Chemoradiation

**DOI:** 10.1101/2024.06.28.24309678

**Authors:** Aaron Seo, Weihong Xiao, Olsi Gjyshi, Kyoko Yoshida-Court, Peng Wei, David Swanson, Tatiana Cisneros Napravnik, Adam Grippin, Aradhana M Venkatesan, Megan C Jacobsen, David T Fuentes, Erica Lynn, Julie Sammouri, Anuja Jhingran, Melissa Joyner, Lilie L Lin, Lauren E Colbert, Maura L Gillison, Ann H Klopp

## Abstract

**Purpose:** The human papillomavirus (HPV) is a significant cause of cervical cancer. We hypothesized that detecting viral cell-free HPV DNA (cfDNA) before, during, and after chemoradiation (chemoRT) could provide insights into disease extent, clinical staging, and treatment response.

**Experimental Design:** Sixty-six patients with locally advanced cervical cancer were enrolled between 2017 and 2023, with 49 receiving standard-of-care (SOC) treatment and 17 participating in a clinical trial combining a therapeutic HPV vaccine (PDS0101; IMMUNOCERV). Plasma samples were collected at baseline, during weeks 1, 3, and 5 of chemoRT, and 3-4 months after chemoRT. HPV cfDNA was quantified using droplet digital PCR targeting the HPV E6/E7 oncogenes of 13 high-risk types. MRI was performed at baseline and before brachytherapy.

**Results:** The median follow-up was 23 months, with recurrence-free survival (RFS) of 78.4% at 2 years. Baseline nodal disease extent correlated with HPV cfDNA levels. HPV cfDNA levels peaked in week 1 of radiation and decreased through treatment. Patients receiving the PDS0101 vaccine had a higher rate of undetectable HPV type 16 cfDNA compared to SOC. HPV cfDNA clearance correlated with better 2-yr RFS (92.9% vs. 30%, log-rank p=0.0067). The strongest predictor of RFS was HPV cfDNA clearance in follow-up achieving a concordance index (CI) 0.83, which improved when combined with MRI response (CI 0.88).

**Conclusions:** HPV cfDNA levels change dynamically during chemoRT. HPV cfDNA levels at first follow-up predict RFS, and the therapeutic HPV vaccine was linked to rapid HPV cfDNA decline. Monitoring HPV cfDNA during and after chemoRT may guide tailoring of personalized treatment.

## INTRODUCTION

Cervical cancer remains a significant health challenge, with a risk of relapse despite aggressive treatment regimens including chemoradiation (chemoRT) with external beam and brachytherapy.^1–3^ Approximately 40% of patients will relapse and 50% will experience some form of chronic toxicity such as diarrhea and pain.^4,5^ These high rates of both relapse and toxicity underscore the urgent need for reliable early markers of relapse to enhance clinical decision-making and patient outcomes.

The role of human papillomavirus (HPV) in the pathogenesis of cervical cancer is well-established, positioning HPV-derived biomarkers as potential candidates for monitoring disease dynamics.^6–8^ Among these, circulating free DNA (cfDNA), including circulating tumor DNA (ctDNA), shows promise in various oncologic settings, reflecting tumor burden and treatment response.^9–12^ However, the detection capabilities and utility of cfDNA in cervical cancer are less well-documented, particularly as an evolving marker over the course of chemoRT, highlighting a gap in our current understanding of how cfDNA clearance can be employed to inform treatment response. The utility of cfDNA as an evolving marker of response to radiation in cervical cancer is unknown. Furthermore, the relative predictive power of cfDNA as compared to radiographic response which is routinely incorporated into cervical cancer treatment has not been evaluated.^13–17^

We proposed that HPV cfDNA can serve as a biomarker for predicting and monitoring treatment response in cervical cancer and may be complementary with imaging-based responses on MRI and ^18^F FDG PET/CT. To evaluate this, we measured HPV cfDNA clearance (a measure of ctDNA in HPV-associated cervical cancers) in patients with cervical cancer receiving definitive chemoRT. Our study incorporates patients treated with either standard-of-care (SOC) chemoRT or with an immunotherapy-chemoRT combination from a recently completed trial involving PDS0101, a novel therapeutic vaccine targeting HPV-related cancers. PDS0101 represents a new avenue in the immunotherapy landscape, and its inclusion in our research allows for a comparative analysis of ctDNA profiles in patients treated with this agent versus SOC.

## MATERIALS AND METHODS

### Study Design: Standard-of-care tissue banking studies

Study patients were enrolled on two longtitudintal tissue banking studies (SWAB2014-0543, CoACH2019-1059). Inclusion criteria are patients ≥ 18 years of age with HPV-positive locally advanced cervical cancer (FIGO 2018 Stages IB3-IVA), who are candidates eligible for curative CRT. Exclusion criteria include those patients < 18 years of age whose tumors are deemed unresectable, those with coexisting medical conditions that would preclude chemotherapy or, radiotherapy, pregnant patients, those unwilling or unable to comply with study and/or follow up procedures and those with contraindications to either contrast enhanced CT or MRI. Treatment for women with locally advanced cervical cancer is concurrent weekly cisplatin (40 mg/m^2^) chemotherapy (up to 5 cycles) with external beam radiotherapy (EBRT) and brachytherapy (BT) delivered in <8 weeks.^18^ Patients received 1.8-2 Gray (Gy) /fraction(fx) of EBRT to a total dose of 45-50 Gy using CT planning followed by brachytherapy (either high dose rate (HDR) or pulsed dose rate (PDR) brachytherapy) and nodal boost if appropriate.

### Study Design: IMMUNOCERV trial protocol

The IMMUNOCERV trial was a single-arm, phase 2 study (NCT04580771; IMMUNOCERV2019-1260) designed to test the hypothesis that PDS0101, a novel, subcutaneously administered HPV-specific vaccine containing peptide pools encoding E6/E7 antigens, would be safe and effective in combination with SOC chemoRT for locally advanced HPV-related cervical cancer. Patients received PDS0101 at Day -10, 7, 28, and 49 (+/-5 days) in relation to the start of SOC chemoRT. Key eligibility criteria included newly diagnosed, biopsy-proven locally advanced squamous cell carcinoma of the cervix (tumor ≥5cm and/or nodal disease), age ≥18 yo, and ECOG ≤2.

### Blood Collection

Peripheral Blood samples were collected in cell-free DNA BCT tubes (Streck, La Vista NE). After centrifuging at 1600 x g for 10 minutes at room temperature, plasma was carefully transferred to microcentrifuge tubes and centrifuged at 16000 x g for 10 minutes to remove any blood cells. Plasma was stored in 1 mL aliquots in a screw cap tubes at -80°C.

### cfDNA isolation

cfDNA was purified with 1 mL of plasma using QIAamp Circulating Nucleic Acid Kit (Qiagen, Hilden, Germany) with vacuum processing following manufacturer’s instruction. Briefly, plasma samples were thawed and digested with proteinase K and lysis buffer containing 1 microgram carry RNA per sample to complete release of nucleic acids from bound proteins, lipids, and vesicles. Lysates were then absorbed onto a QIAamp Mini column by vacuum pressure. After washing the column, circulating nucleic acids were eluted in 60 μl of AVE buffer and stored at -20°C until analysis.

### cfHPV ddPCR analysis

To detect high-risk single copies of HPV16 or HPV18, 31, 33, 35, 39, 45, 51, 52, 56, 58, 59 and 68 DNA, a dual-target droplet digital PCR (ddPCR) method was developed using the sequence of their HPV E6 or E7 oncogenes.^19^ The specific primers and probes for 7 dual-target PCR were designed including HPV sequence variance and phylogenetic variables, the probes were differentially fluorescence-tagged for the simultaneous detection and quantification of both types. Human endogenous retrovirus group 3 (ERV3) gene was used as human DNA quality control. Seven dual-target ddPCR assays are pairs of HPV16-ERV3, HPV18-HPV33, HPV33-HPV35, HPV31-HPV58, HPV52-HPV59, HPV39-HPV51 and HPV56-HPV68. The primers and probes for each HPV type were designed to avoid cross reaction among 13 high risk HPV types. HPV ddPCR assay were performed on the QX200 ddPCR System (Bio-Rad, Hercules, CA). Each reaction assay contained 5 μl of 4x ddPCR supermix for probe (no dUTP), 0.9 nM of respective primers, 0.25 nM of respective probes, 4 units of restriction enzyme BamH1 and 13 μl of elute in a final volume of 20 μl. The mix were vortexed and loaded into Bio-Rad carriage to generate the droplets, 40 μl of droplets were transfer slowly into the PCR plate and sealed with pierceable foil. PCR was performed as 1 cycle of 95°C for 10 minutes, 40 cycles of 94°C for 30 seconds and 57.1°C for 1 minute, 1 cycle of 95°C for 10 minutes, and all step ramp rate was 2°C per second. The amplified droplets were detected in Bio-rad Reader. Data were analyzed to determine HPV types and copy numbers using QuantaSoft software. Measures of ≥16 copies/mL considered quantifiable were previously described and were described as detectable for purposes of the analysis.^19^ “Undetectable” and “detectable” cfDNA were designated “cleared” and “uncleared” statuses, respectively.

### Statistical Analysis

The chi-squared test was used to compare the distributions of HPV types between primary tumor swab and peripheral blood. cfDNA levels below 16 were adjusted to the quantifiable threshold of 16 copies/mL. Then, all values underwent log10-transformation for further analysis. The Fisher’s Exact test and Wilcoxon rank sum test were used to compare sociodemographic and clinical factors across cohort types. The Shapiro-Wilk test was used for normality testing of continuous variables. The Kruskal-Wallis test was used to associate cfDNA levels at different timepoints with sociodemographic and clinical factors. Dunn’s test was used to conduct *post hoc* pairwise comparisons to identify specific groups with differences identified by the Kruskal-Wallis test.

Recurrence-free survival (RFS) was defined as the time from histological diagnosis of cervical cancer to the time of first recurrence. The Kaplan-Meier method was used to estimate RFS and compared with the log-rank test. Factors associated with RFS were evaluated with the Cox proportional hazards model. Univariable analysis included the following factors: cohort designation, age at diagnosis, ethnicity, body mass index, smoking status, FIGO 2018 stage, highest involved lymph node at diagnosis, number of cisplatin cycles, baseline MRI gross tumor volume (GTV), MRI GTV during week 5 of treatment, GTV regression (defined as: 1-(GTV_Week 5 of treatment_/GTV_Baseline_. Optimal GTV regression was defined as GTV regression ≥0.90 as previously described),^13^ HPV cfDNA levels before treatment, during week 1 of treatment, during week 3 of treatment, during week 5 of treatment, and in follow-up, 3-4 months after completion of therapy. Given the limited follow-up sample size and events, multivariable analysis for RFS was restricted to two variables: follow-up HPV cfDNA level (continuous) and number of cisplatin cycles.

Concordance indices (c-indices) were calculated to quantify the predictive accuracy of predictive variables. C-index is analogous to the area under the receiver operating characteristic curve (AUCROC) for binary outcomes but modified to account for censoring in time-to-event outcomes. Like AUCROC, a c-index of 1.0 indicates perfect classification whereas a value of 0.5 suggests random guessing. All statistical analyses were performed using R version 4.2.1.

## RESULTS

### Cohort Demographic and Clinical Characteristics

Between 2017 and 2023, 66 patients with locally advanced cervical cancer were enrolled on this longitudinal study (49 SOC patients, 17 PDS0101 patients). All patients were treated with definitive chemoRT with brachytherapy boost, and all patients received concurrent cisplatin. Patients on the PDS0101 study received the HPV-directed vaccine before, during, and after chemoRT. For the overall cohort, the median follow-up was 23 months, with a 2-year RFS of 78.4% (95% confidence interval (CI) 67.5%-91.0%).

Sociodemographic factors were balanced between the two cohorts (**Table 1**). Clinical factors such as FIGO 2018 Stage, baseline and mid-treatment GTV MRI volume measurements, GTV treatment response on MRI, number of cisplatin cycles, baseline and follow-up maximum SUV on ^18^F FDG PET/CT were balanced between the two cohorts (**Table 2**). The pattern of lymph node involvement between the PDS0101 and SOC cohorts was different, with greater lymph node involvement overall in PDS0101 compared to SOC (100% vs. 73%), though PDS0101 patients had predominantly pelvic lymph nodes as the highest clinically involved lymph node level, whereas SOC patients were more likely to have common iliac and para-aortic lymph node involvement (P=0.009) (**Table 2**).

**Table 1.** Sociodemographic Characteristics.

| Characteristic | Overall, N = 66<sup>1</sup> | PDS, N = 17<sup>1</sup> | SOC, N = 49<sup>1</sup> | p-value<sup>2</sup> |
| --- | --- | --- | --- | --- |
| Age (yrs) | 44.8 (11.7) | 44.1 (14.2) | 45.0 (10.9) | 0.5 |
| BMI (kg/m^2) | 28.9 (5.9) | 26.7 (3.7) | 29.7 (6.4) | 0.12 |
| Ethnicity |  |  |  | 0.2 |
| Declined to answer | 2 (3.0%) | 1 (5.9%) | 1 (2.0%) |  |
| Hispanic or Latino | 18 (27%) | 2 (12%) | 16 (33%) |  |
| Not Hispanic or Latino | 46 (70%) | 14 (82%) | 32 (65%) |  |
| Smoking Status |  |  |  | 0.8 |
| Current | 8 (12%) | 2 (12%) | 6 (12%) |  |
| Former | 19 (29%) | 6 (35%) | 13 (27%) |  |
| Never | 39 (59%) | 9 (53%) | 30 (61%) |  |

<sup>1</sup> Mean (SD); n (%)
<sup>2</sup> Wilcoxon rank sum test; Fisher's exact test

**Table 2.** Clinical Characteristics. Abbreviations: FIGO, International Federation of Gynecology and Obstetrics; GTV, gross tumor volume; EBRT, external beam radiotherapy; SUV, standardized uptake values; PET-CT, positron emission tomography computed tomography.

| Characteristic | Overall, N = 66<sup>1</sup> | PDS, N = 17<sup>1</sup> | SOC, N = 49<sup>1</sup> | p-value<sup>2</sup> |
| --- | --- | --- | --- | --- |
| FIGO 2018 Stage |  |  |  | 0.6 |
| I | 4 (6.1%) | 1 (5.9%) | 3 (6.1%) |  |
| II | 12 (18%) | 2 (12%) | 10 (20%) |  |
| III | 46 (70%) | 12 (71%) | 34 (69%) |  |
| IV | 4 (6.1%) | 2 (12%) | 2 (4.1%) |  |
| Highest Node Involved Node |  |  |  | 0.009 |
| Common_Iliac | 12 (18%) | 2 (12%) | 10 (20%) |  |
| Not_Involved | 13 (20%) | 0 (0%) | 13 (27%) |  |
| Para-Aortic | 12 (18%) | 2 (12%) | 10 (20%) |  |
| Pelvic | 29 (44%) | 13 (76%) | 16 (33%) |  |
| Baseline GTV Volume on MRI (cm^3) | 71 (78) | 68 (59) | 72 (84) | 0.5 |
| Mid-EBRT GTV Volume on MRI (cm^3) | 14 (28) | 13 (20) | 14 (30) | 0.2 |
| Unknown | 6 | 0 | 6 |  |
| GTV Treatment Response on MRI |  |  |  | 0.11 |
| Optimal | 27 (46%) | 5 (29%) | 22 (52%) |  |
| Sub-Optimal | 32 (54%) | 12 (71%) | 20 (48%) |  |
| Unknown | 7 | 0 | 7 |  |
| Cisplatin Cycles Received |  |  |  | 0.6 |
| 1-3 | 4 (6.1%) | 2 (12%) | 2 (4.1%) |  |
| 4+ | 58 (88%) | 14 (82%) | 44 (90%) |  |
| Other | 4 (6.1%) | 1 (5.9%) | 3 (6.1%) |  |
| Baseline Maximum SUV on PET-CT | 19 (13) | 19 (7) | 19 (15) | 0.4 |
| Unknown | 4 | 1 | 3 |  |
| 3-4 mo f/u Maximum SUV on PET-CT | 3.47 (1.66) | 2.96 (1.21) | 3.69 (1.79) | 0.063 |
| Unknown | 9 | 0 | 9 |  |

<sup>1</sup> n (%); Mean (SD)
<sup>2</sup> Fisher's exact test; Wilcoxon rank sum test; Pearson's Chi-squared test

### Baseline HPV Tumor and Plasma Genotyping

HPV typing of primary tumor swabs showed that 70% (35/50) of tumors were HPV type 16 or 18, 16% (8/50) were other high-risk HPV types, and 14% (7/50) were negative (**Figure 1A**). The distributions of HPV types in cfDNA compared to tumor swabs was similar (Chi-squared test, p=0.99), with 77% (51/66) of cfDNA HPV types were 16 or 18, 14% (9/66) were other high-risk HPV types, and 9.0% (6/66) were negative (**Figure 1B)**. One patient with primary tumor swab HPV type 16 was negative in cfDNA, suggesting a non-shedding status. Two patients had discordant positive HPV types in primary tumor swab and plasma (one patient with type 31 in tumor and type 18 in plasma; one patient with type 16 in tumor and type 45 in plasma). Of the 7 patients with no HPV type detected in the primary tumor with swab testing, 4 patients had a detectable high-risk HPV type in plasma (2 patients with HPV type 16 and 2 patients with HPV type 18).

**Figure 1.**
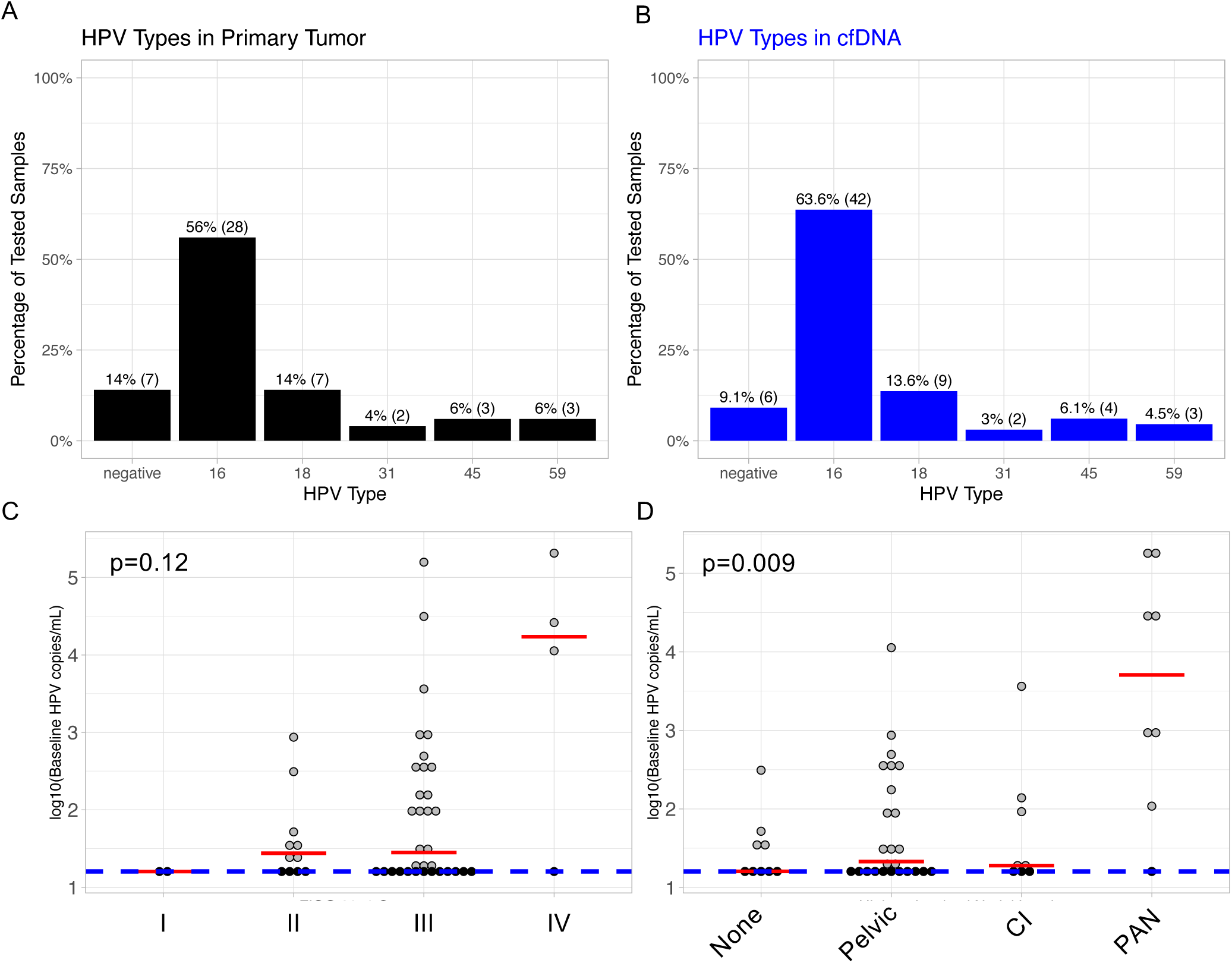
HPV types and baseline characteristics. (A) HPV types from primary tumor swab and (B) HPV types from cfDNA for patients from whom both tumor swab and cfDNA HPV typing was performed. (C) Pre-treatment cfDNA levels by FIGO 2018 Stage. (D) Pre-treatment cfDNA levels by the highest involved lymph node at diagnosis. Grey dots: detectable cfDNA levels; black dots: below limit of detection; Red line: the median value; blue dotted line: limit of detection. Kruskal-Wallis rank sum p-values are reported.

### Baseline HPV cfDNA Levels and Clinical Characteristics

With the 50 available baseline HPV cfDNA levels, the median value was 27.9 copies/mL (range: 16.0 – 2.06 × 10^5^). The differences in baseline HPV cfDNA levels across FIGO 2018 stage were not statistically significant (p=0.11, **Figure 1C**), though Stage IV patients had the highest median value (1.87 × 10^4^ copies/mL) compared to Stages I, II, and III (16.0, 27.6, 28.2, respectively). Baseline HPV cfDNA levels were significantly different across different categories of the highest echelon involved lymph node level at time of diagnosis (p=0.02, **Figure 1D**). Patients with para-aortic involvement had the highest median baseline HPV cfDNA (1.36 × 10^4^ copies/mL), which on *post hoc* pairwise comparisons was significantly different compared to common iliac involvement (median: 19.0 copies/mL, p=0.0076), pelvic lymph node involvement (median: 21.4 copies/mL p=0.0012), and having no involved lymph nodes (median: 16.0 copies/mL, p=0.0011) (**Figure 1D**). There was no significant difference in *post hoc* pairwise comparisons of baseline HPV cfDNA levels in other levels of lymph node involvement. Baseline cfDNA levels were not significantly correlated to baseline GTV on MRI (p=0.24) or baseline maximum SUV on ^18^F FDG PET/CT (p=0.57).

### HPV cfDNA Kinetics During and After Definitive ChemoRT

During treatment, the median HPV cfDNA level peaked during Week 1 (median: 42.3 copies/mL, range: 16.0 - 3.13 × 10^4^) and subsequently decreased through treatment (Week 3: median 16.0 copies/mL, range 16.0 – 1.25 × 10^5^; Week 5: median 16.0 copies/mL, range 16.0 – 3,780) and in 3-4 month follow-up (median 16.0 copies/mL, range 16.0 – 789) (**Figure 2A**). Of the 66 patients with at least one timepoint measurement available, 6 patients (9.1%) never had cfDNA detected (**Figure 2A, Supplementary Figure 1**). Of the 42 patients with at least three timepoint measurements, 31% of patients had peak cfDNA levels at baseline, and 50% of patients had peak cfDNA levels during Week 1 of treatment (**Figure 2B**). Clearance of cfDNA levels progressively increased during treatment, starting with HPV cfDNA undetected at baseline in 40% of patients, HPV cfDNA undetected during Week 1 of treatment in 35.6% of patients, HPV cfDNA undetected during Week 3 of treatment in 60.9% of patients, and HPV cfDNA undetected during Week 5 of treatment in 81.0% of patients (**Supplementary Figure 2A**). 76.2% of patients had undetected HPV cfDNA at 3-4 month follow-up.

**Figure 2.**
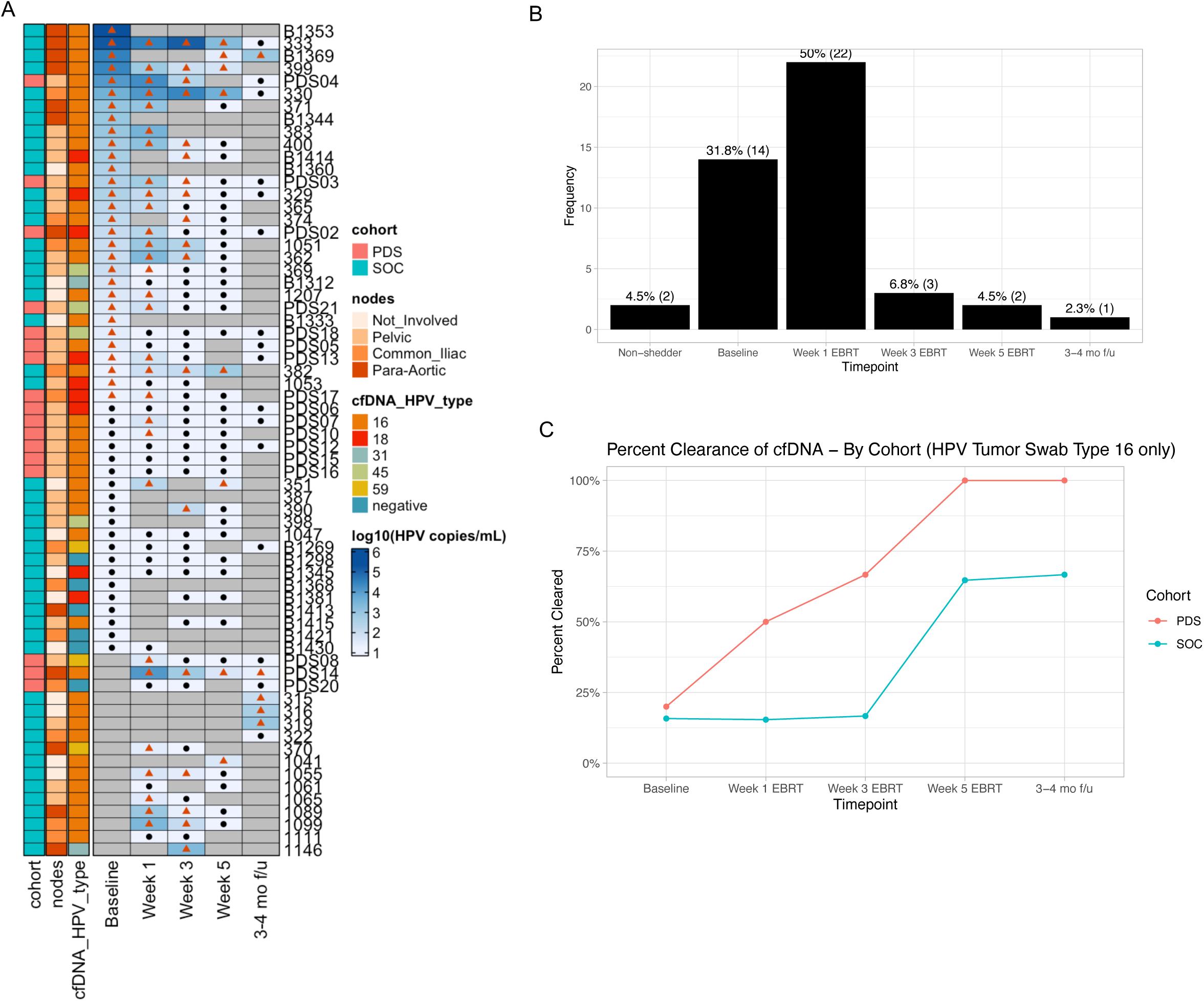
HPV cfDNA Kinetics. (A) Heatmap of HPV cfDNA levels from before, during, and after definitive chemoradiation. Grey shade: Not tested; black dot: level below limit of detection; red square: above limit of detection (B) Counts of the timepoints at which peak cfDNA levels were measured in patients with at least three timepoint measurements. (C) Percentage of patients with HPV cfDNA type 16 clearance (below limit of detection) by timepoint.

### Earlier HPV type 16 cfDNA Clearance in Patients Receiving PDS0101 Therapeutic Vaccine Compared to SOC

Because the PDS0101 vaccine is derived from HPV type 16 peptides, we hypothesized that patients who received the PDS0101 vaccine would have greater HPV type 16 cfDNA clearance rate compared to SOC. The PDS0101 vaccine cohort had a greater percentage of patients with undetectable HPV type 16 cfDNA at each ascertainable timepoint (**Figure 2C).** Furthermore, patients who received the PDS0101 vaccine also experienced greater HPV cfDNA clearance at each timepoint when considering all HPV types. (**Supplementary Figure 2B**).

### Detected HPV cfDNA at 3-4 Month Follow-up Associated with Reduced Recurrence-free Survival

Given that HPV cfDNA clearance increases throughout treatment and plateaus near the end of treatment, we hypothesized that the presence of HPV cfDNA near the end of treatment and at 3-4 month follow-up would be associated with inferior survival outcomes. HPV cfDNA clearance at baseline or during treatment (Weeks 1, 3, and 5) were not significantly associated with RFS (**Figure 3A-C**). Detectable vs. undetectable HPV cfDNA at 3-4 month follow-up were associated with worse RFS (2-year RFS estimate 30.0% [95% CI 6.31%-100%] vs. 92.9% [95% CI 80.3%-100%], log-rank p=0.0067). (**Figure 3D)**. On univariable Cox regression analysis, HPV cfDNA levels at 3-4 month follow-up was the only significant variable associated with RFS (HR 7.21 [95% CI 1.79-28.96, p=0.005] (**Table 3)**.

**Figure 3.**
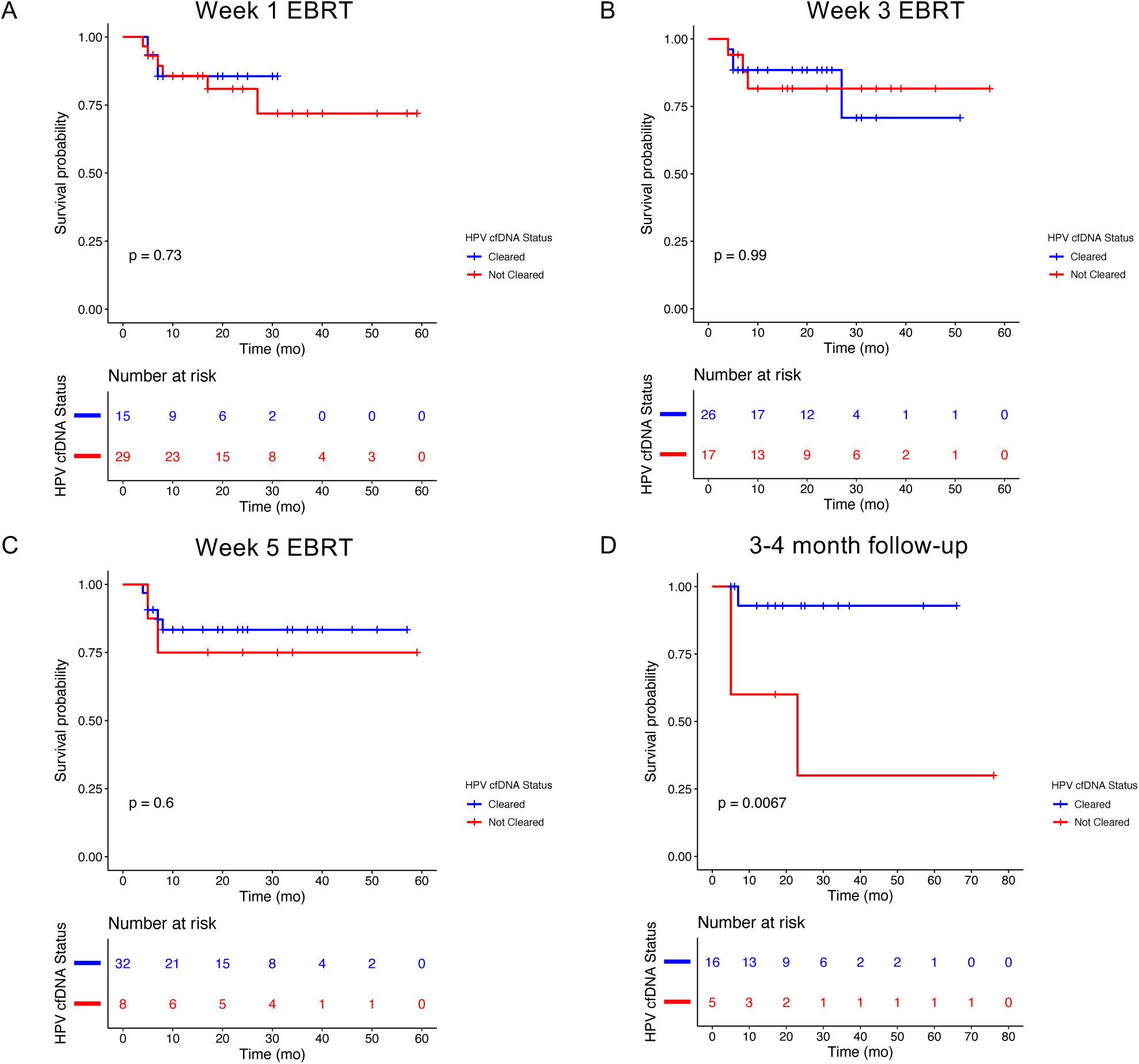
Kaplan-Meier curves for recurrence-free survival stratified by detectable vs. undetectable HPV cfDNA (A) during Week 1 EBRT, (B) during Week 3 EBRT, (C) During Week 5 EBRT, and (D) at 3-4 mo follow-up. The logrank test p-values are reported.

**Table 3.**
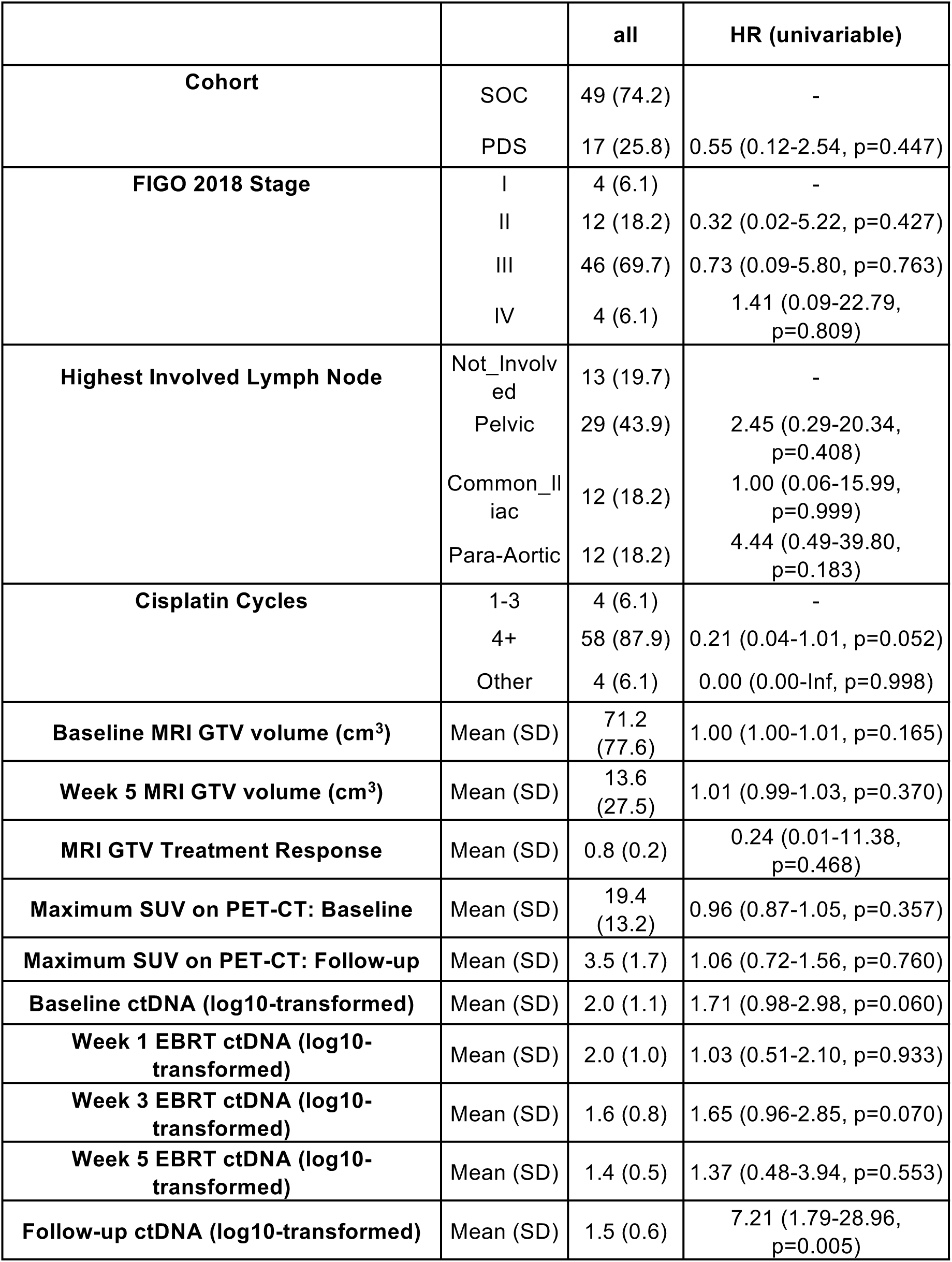
Univariable Cox Regression Analysis for Recurrence-Free Survival.

### Predictive Accuracy of HPV cfDNA levels, MRI, and ^18^F FDG PET/CT Responses on Recurrence-free Survival

To compare the performance of HPV cfDNA levels and various imaging modalities to predict RFS and to determine if these variables were complementary or synergistic, we measured the concordance indices (c-indices) for HPV cfDNA levels at 3-4 month follow-up, GTV volumetric reduction from before treatment to pre-brachytherapy (MRI response), and PET/CT FDG-avidity at 3-4 month follow-up (PET/CT). C-indices for HPV cfDNA levels at 3-4 month follow-up, MRI response, and ^18^F FDG PET/CT (maximum SUV at follow-up) were 0.83 ± 0.12, 0.60 ± 0.09, and 0.49 ± 0.11, respectively. The c-index for HPV cfDNA levels at 3-4 month follow-up and MRI response combined was 0.88 ± 0.1. Multivariable Cox regression analysis showed an increased hazard ratio for HPV cfDNA levels at 3-4 month follow-up when adjusted for MRI (HR 9.54 [95% CI 0.920-99.0, p=0.0588]) (**Supplementary Table 2**).

## DISCUSSION

In this prospective collection study from a large tertiary cancer center, we describe HPV cfDNA kinetics before, during, and after definitive chemoRT in patients with locally advanced cervical cancer, including patients receiving the PDS0101 therapeutic vaccine derived from HPV E6 and E7 peptides. In the pooled analysis, quantifiable HPV cfDNA at 3-4 month follow-up was associated with inferior RFS, whereas HPV cfDNA levels at other timepoints before and during chemoRT were not associated with RFS.

HPV types in primary tumor and plasma were largely concordant, with most of the discordances stemming from undetected HPV types in primary tumor with detection in plasma. One potential explanation for this discordant pattern is a false negative primary tumor swab from insufficient sampling, where plasma HPV cfDNA could assist in accurately characterizing the HPV association of a patient’s cervical cancer, which can have important prognostic implications as HPV-negative cervical cancers are associated with worse outcomes than those that are HPV-associated.^20–25^

Baseline HPV cfDNA levels were most strongly associated with the highest involved lymph node at time of diagnosis, with the highest levels in patients with para-aortic lymph node involvement. Of note, primary tumor size was not associated with baseline HPV cfDNA level, suggesting that circulating tumor DNA is most reflective of regional spread rather than local disease burden.

We show that HPV cfDNA levels generally rise and peak early during chemoRT, suggestive of tumor cellular death and genomic shedding early in treatment. HPV cfDNA levels subsequently nadir at the completion of chemoRT. The slight increase in percent HPV cfDNA clearance at 3-4 month follow-up suggests HPV cfDNA clearance can continue to occur after completion of definitive chemoRT.

Of note, on exploratory sub-analysis, the patients who received PDS0101, a therapeutic vaccine derived from HPV type 16 E6 and E7 peptides, experienced greater and earlier clearance of HPV type 16 cfDNA compared to patients receiving SOC treatment. This difference could reflect vaccine efficacy with respect to immune system recognition of the E6 and E7 viral peptides. Additional detailed immunoprofiling from patient samples will help clarify the effects of the therapeutic vaccine. Though patients receiving PDS0101 were included in this report, the study was not statistically powered for survival outcomes comparisons between cohorts, which will be the focus of additional studies.

A select number of studies have explored serial measurements of HPV cfDNA levels.^26–33^ The largest prospective study to date assessing ctDNA levels in cervical cancer patients receiving definitive chemoRT (Han *et al.)* evaluated 70 total patients at three timepoints: end of chemoRT, 4-6 weeks post-chemoRT, and 3 months post-chemoRT.^34^ Our study provides unique insights into early changes in HPV cfDNA during radiation treatment. We observed an increase in HPV cfDNA for patients during the first week of radiation. This rapid increase in tumor DNA in circulation may reflect intracellular contents being released upon cell death. Although we did not detect a relationship between week 1 cfDNA and outcome, these early changes suggest that rapid cfDNA kinetic changes may be a potential as an early response biomarker, meriting further evaluation with additional time points and patients.

Han *et al.* found that detectable HPV ctDNA at all three timepoints were significantly associated with inferior 2-year progression-free survival (PFS) with a median lead time to recurrence of 5.9 months. In contrast, our study did not find a significant difference in RFS with quantifiable HPV cfDNA in the last week of chemoRT. This difference could be attributed to our study having too few samples in the later timepoints for analysis. Han *et al.* utilized both ddPCR and a next-generation sequencing approach and found that both approaches had similar results. This finding lends credence to our use of ddPCR and also has encouraging implications for testing access worldwide, as ddPCR is, in general, less costly than next-generation sequencing technologies.^35^ Furthermore, the performance of HPV cfDNA levels at 3 month follow-up to predict RFS/PFS based on c-index was equivalent in our study and Han *et al.* (c-index = 0.72).^34^ Though Han *et al*. did not include routine ^18^F FDG PET/CT follow-up as part of their study, our study showed that combining HPV cfDNA levels and ^18^F FDG PET/CT follow-up yields a higher c-index (0.92), suggesting a multimodal surveillance approach may be more effective at detecting early recurrence. Unlike Han et al, our study evaluated the combined use of on-treatment MRI response and cfDNA response and suggested an additive benefit of these two biomarkers. Larger prospective cohorts will help validate these results.

Although prior randomized trial efforts to identify a patient population who would benefit from “outback” chemotherapy have failed, our study suggests that HPV ctDNA levels after chemoRT can help identify a population at higher risk of relapse who may benefit from treatment escalation with adjuvant systemic therapy.^36^ Additionally, in light of conflicting evidence from recently reported phase 3 clinical trials utilizing concurrent and adjuvant immunotherapy,^37,38^ intra-treatment ctDNA monitoring may help identify patients with excellent early response who may benefit from either treatment escalation or treatment de-escalation from a combined IO-chemoRT to avoid toxicities associated with systemic immunotherapy.

The strengths of this study include robust imaging correlates including primary tumor HPV swab typing, cfDNA monitoring early in radiation treatment, mid-treatment MRI and 3-4 month follow-up ^18^F FDG PET/CT scan in the majority of patients, use of modern radiation therapy techniques, use of Streck tubes for optimal plasma preservation, multiple intra-treatment timepoints, and the first report of ctDNA kinetics in patients receiving the PDS0101 therapeutic vaccine. Limitations of the study include limited follow-up sample collection and no additional timepoints between end of chemoRT and 3-4 month follow-up.

In conclusion, in this study, quantifiable HPV cfDNA at 3-4 month follow-up was associated with inferior RFS. The PDS0101 therapeutic vaccine was associated with greater and earlier clearance of HPV type 16 cfDNA. Additional follow-up and studies are needed to assess for differences in survival outcomes with PDS0101 and to ascertain the clinical utility of HPV cfDNA testing during and after chemoRT to guide treatment de-escalation as well as escalation with adjuvant systemic therapy.

## AUTHOR CONTRIBUTIONS

The study was conceived and designed by AS, OG, MLG, and AHK. Clinical data collection was performed by AS, OG, KY-C, TCN, MCJ, DTF, EL, and JL. ddPCR assay was performed by WX. Data analysis and statistics were done by AS. AS drafted the manuscript. AS, WX, OG, KY-C, PW, DS, TCN, AG, AMV, MCJ, DTF, EL, JS, AJ, MJ, LLL, LEC, MLG, and AHK edited the manuscript. All authors had the opportunity to review the manuscript and agree with the enclosed contents.

## ACKNOWLEDGEMENTS

We would like to thank Van Morris and Jia Sun for helpful discussions.

## ETHICS STATEMENT

The clinical studies were conducted according to the guidelines of the Declaration of Helsinki and approved by the Institutional Review Board of the University of Texas MD Anderson Cancer Center (SWAB2014-0543, CoACH2019-1059, IMMUNOCERV2019-1260). Informed consent was obtained from all study participants.

## CONFLICT OF INTEREST STATEMENT

Authors declare no conflict of interest.

## DATA AVAILABILITY STATEMENT

The de-identified data that support the findings of this study will be available on reasonable request. The data are not publicly available due to protected health information that could compromise the privacy of research participants.

## Notes

### Competing Interest Statement

The authors have declared no competing interest.

### Funding Statement

This study was funded by the University of Texas MD Anderson Cancer Center Human Papillomavirus Moonshot Program.

### Author Declarations

The Institutional Review Board at the University of Texas MD Anderson Cancer center gave ethical approval for this work.

