## Supplementary Information for "Human Papilloma Virus Circulating Cell-Free DNA Kinetics in Cervical Cancer Patients Undergoing Definitive Chemoradiation"

### Supplementary Figures and Tables

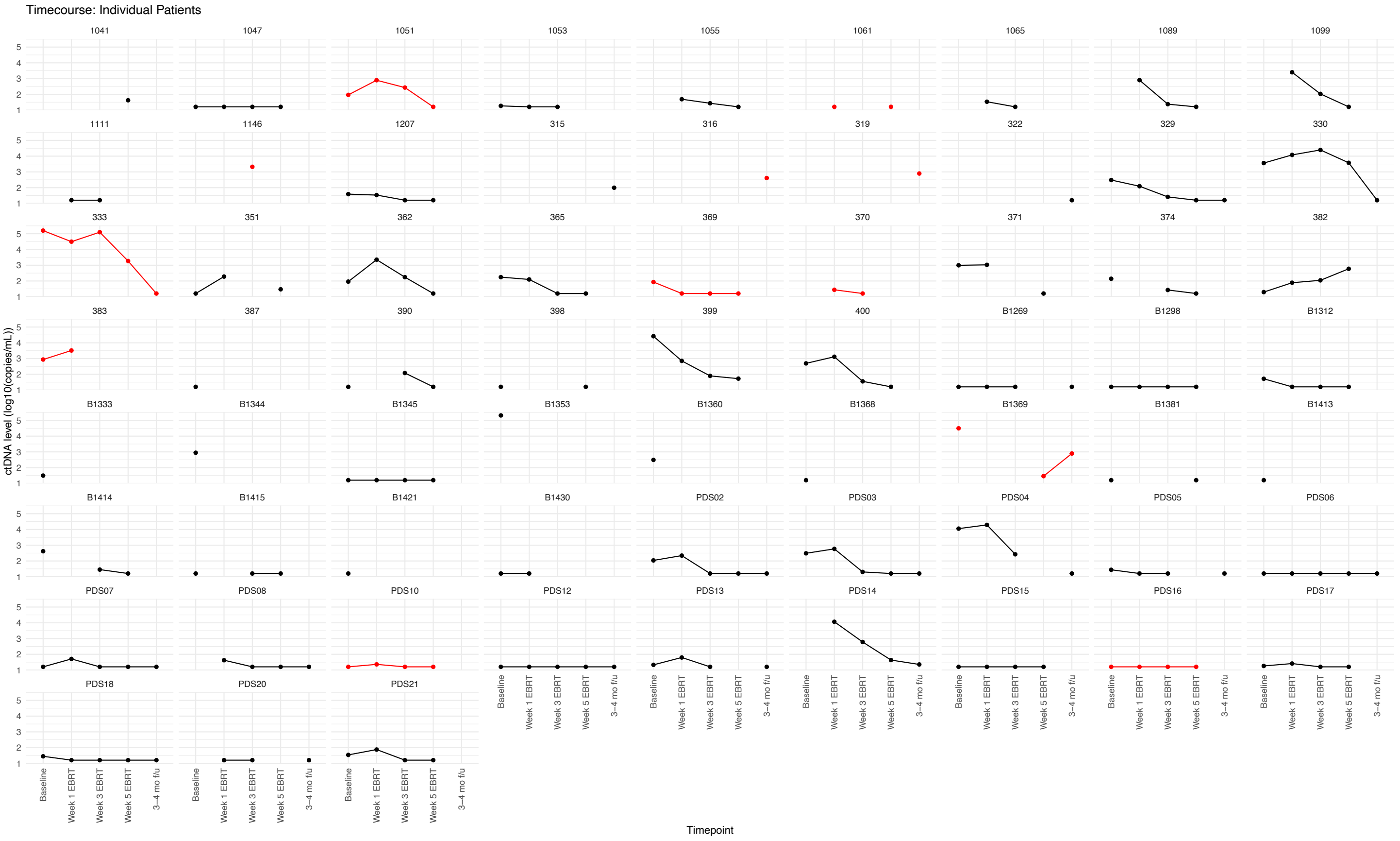


**Supplementary Figure 1.** Individual timecourse plots of ctDNA levels before, during, and after chemoRT. Red indicates patients who had an eventual recurrence.

A

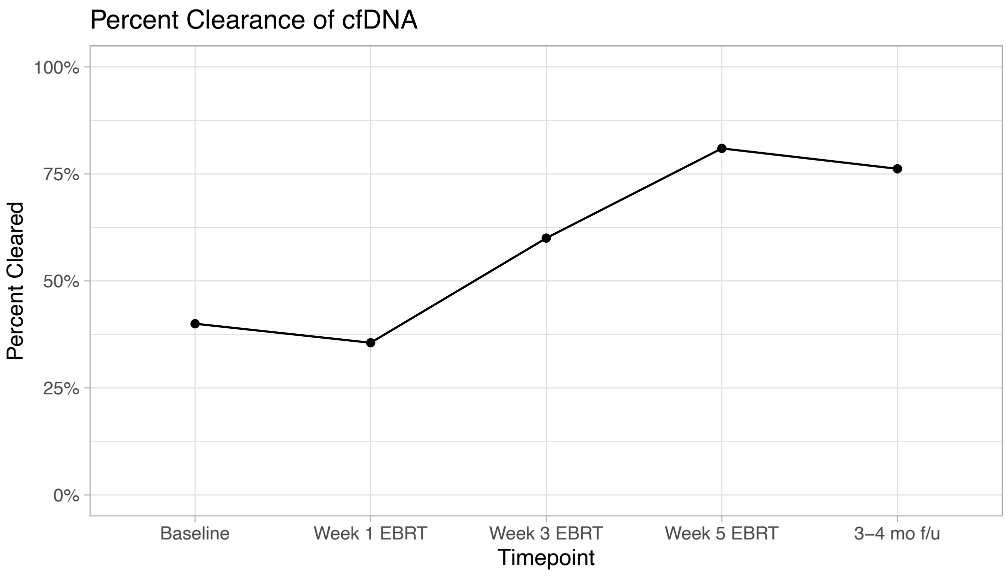


B

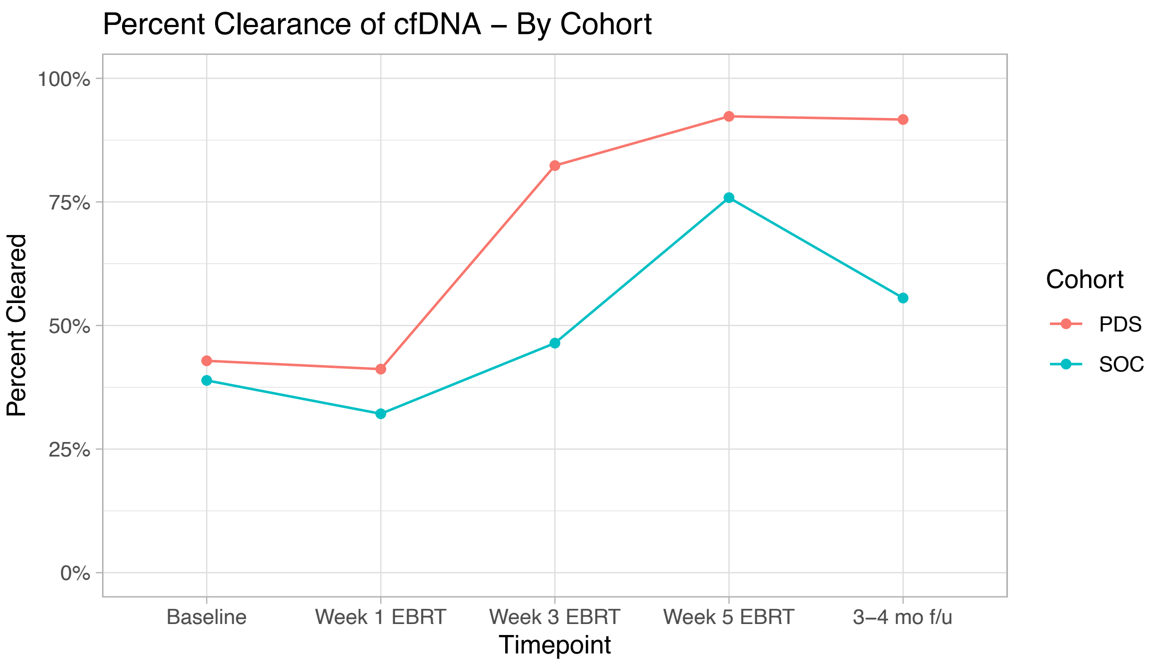


**Supplementary Figure 2**. Percent clearance of HPV cfDNA of all HPV types (A) overall and (B) by cohort.

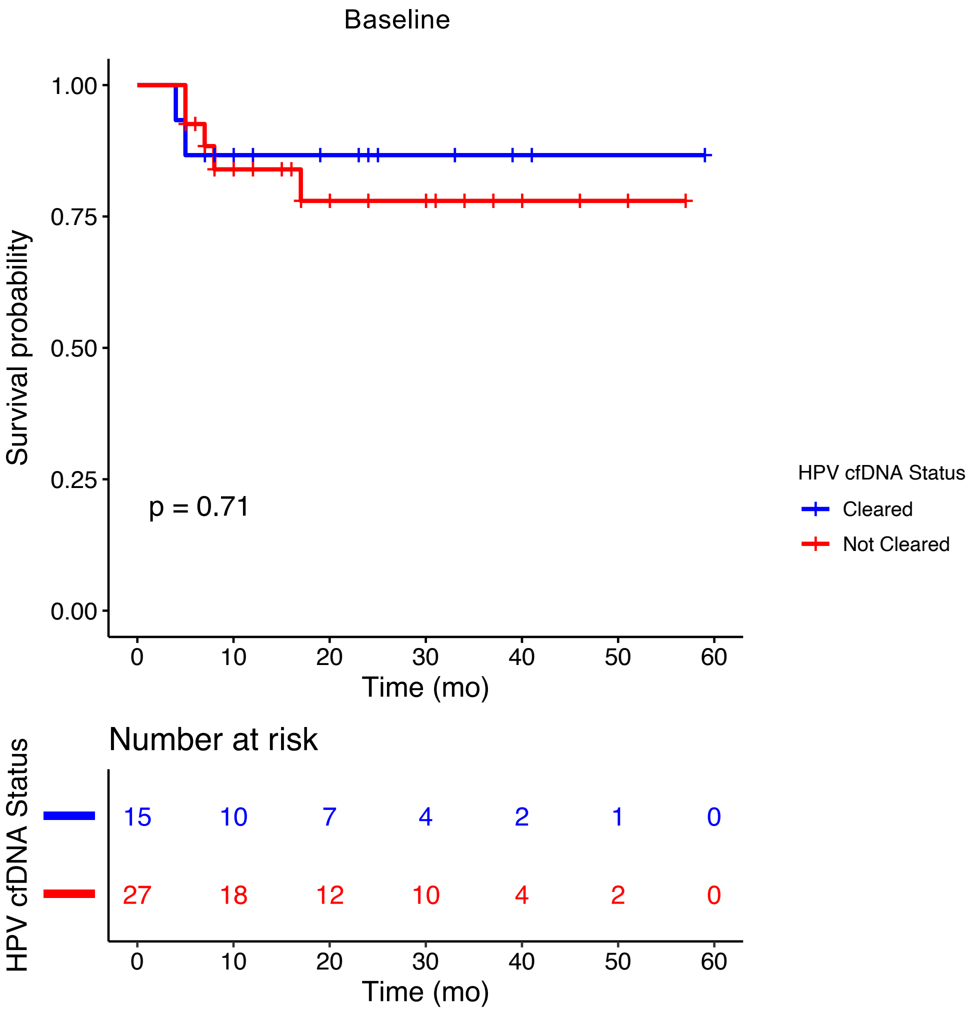


**Supplementary Figure 3.** Kaplan-Meier Analysis of Recurrence-Free Survival by Baseline ctDNA status. The logrank test p-value is reported.

| | Univariable | | Multivariable | |
|---|---|---|---|---|
| **Variable** | **HR (95% CI, p-value)** | **C-index** | **HR (95% CI, p-value)** | **C-index** |
| **Follow-up ctDNA (log10-transformed)** | 7.21 (1.79-29.0, p=0.0054) | 0.828 ± 0.116 | 9.54 (0.920-99.0, p=0.0588) | 0.881 ± 0.098 |
| **MRI GTV treatment response** | 0.240 (0.00505-11.4, p=0.468) | 0.601 ± 0.09 | 0.0104 (1.81x10$^{-9}$-60016, p=0.566) | |

**Supplementary Table 2.** Univariable and Multivariable Cox Regression Analyses and C-indices for Recurrence-free Survival.
